# Identifying Indicators of Advanced Levels of Clinical Competence in Musculoskeletal Physiotherapy: A Francophone Delphi Study

**DOI:** 10.64898/2026.09.29.26363758

**Authors:** Adrien Mounier-Poulat, Nicolas Pinsault, Romain Artico

## Abstract

**Background:** As musculoskeletal physiotherapy evolves toward advanced practice and direct access, evaluating high levels of clinical competence has become pressing. Quality indicators underpin large-scale audit systems, but indicators discriminating advanced competence remain insufficiently defined.

**Objective:** To identify and characterize indicators capable of discriminating high levels of clinical competence in musculoskeletal physiotherapy.

**Methods:** A modified Delphi comprised an exploratory pre-round and two consensus rounds. Experienced French-speaking musculoskeletal physiotherapists were recruited using an expertise scoring grid. Candidate indicators, generated from open-ended responses and existing quality indicators, were rated on 6-point Likert scales for relevance, formulation, and tool adequacy. Consensus was predefined as ≥75% of ratings ≥5.

**Results:** Thirty-two experts completed the pre-round, 29 the first round, and 27 the second (84% retention), generating 49 candidate indicators. Thirteen reached consensus for relevance, and 10 also for formulation. Retained indicators primarily reflected advanced clinical reasoning: real-time adaptation, hypothesis prioritization, decision-making under uncertainty, and error management. Only one outcome indicator reached consensus: clinical performance weighted by case complexity.

**Conclusion:** High levels of clinical competence in musculoskeletal physiotherapy appear characterized less by standardized outcome attainment than by managing complexity, uncertainty, and error through advanced clinical reasoning. These findings may inform competence-oriented audit and feedback systems.

**Trials registration:** Not applicable.

## INTRODUCTION

Musculoskeletal disorders represent the leading cause of years lived with disability worldwide, accounting for more than 149 million years lived with disability in 2020, with projections indicating continued growth by 2050 [1]. Their health, economic, and societal burden positions musculoskeletal physiotherapy as a central component of primary care systems, where most management episodes and long-term care trajectories occur. Beyond access, contemporary challenges involve chronicity, multimorbidity, and the management of complex, evolving situations within constrained organizational environments [2]. In this context, defining and evaluating high levels of clinical competence has become critical.

Healthcare systems traditionally rely on quality indicators to monitor and improve care. Grounded in Donabedian’s structure–process–outcome model [3], these indicators assess adherence to standards, safety, and overall performance at organizational or population levels [4-7]. They underpin public quality policies and audit systems and have demonstrated effectiveness in improving healthcare performance [8-12].

However, evaluating clinical practice requires distinguishing three related but non-equivalent constructs: clinical competence, performance, and quality of care [13,14]. Clinical competence refers to a practitioner’s relatively stable capacity to mobilize cognitive, technical, relational, and contextual resources to manage singular situations characterized by uncertainty and evolving complexity [15-17]. Performance is the situated expression of that competence in real practice, shaped by individual and system-related influences including patient characteristics, case complexity, and organizational conditions [14]. Quality of care, in contrast, is a systemic property concerned with safety, conformity, and efficiency at organizational or population levels, largely determined by factors external to the individual clinician [3]. Competence is therefore expressed through situated clinical reasoning and adaptive decision-making rather than through outcome attainment alone [18].

This distinction has direct implications for evaluation. Outcomes such as pain reduction, functional improvement, recurrence, or patient satisfaction measure performance rather than competence: they are influenced by biopsychosocial determinants, case complexity, and organizational constraints beyond the practitioner’s control [19,20]. While essential for accountability, their capacity to differentiate highly competent practitioners from those meeting baseline standards remains limited.

Audit and feedback research indicates that performance monitoring is particularly effective in improving lower-performing clinicians [8-11], but its impact is more modest among experienced practitioners. When indicators target compliance or global outcomes, they may insufficiently capture higher-order cognitive processes associated with advanced levels of clinical competence.

This limitation is especially salient in musculoskeletal physiotherapy. Recent systematic reviews have identified robust quality indicators for primary and community musculoskeletal care [21], yet these do not explicitly differentiate levels of clinical competence and remain predominantly population oriented.

The literature on expertise and deliberate practice emphasizes that advanced levels of competence develop through sustained, targeted feedback on reasoning and decision-making in complex situations [19,22,23]. Advanced levels of clinical competence may therefore be characterized by the ability to manage diagnostic uncertainty, prioritize competing hypotheses, anticipate non-linear trajectories, integrate biopsychosocial determinants into prognostic reasoning, and adapt management over time—dimensions not easily operationalized within conventional quality frameworks.

International rehabilitation competency frameworks similarly call for clearer markers of advanced clinical practice [24,25]. To date, however, no study has specifically sought to identify indicators designed to discriminate advanced levels of clinical competence in musculoskeletal physiotherapy.

To address this gap, the present study aimed to identify and characterize indicators capable of discriminating advanced levels of clinical competence in musculoskeletal physiotherapy, with a view to supporting more targeted, competence-oriented evaluation systems. Advanced clinical competence was defined as the capacity to mobilize refined clinical reasoning, manage uncertainty and case complexity, ensure patient safety, and optimize care trajectories.

## METHODS

### Study design

This study used a modified Delphi methodology comprising an exploratory pre-round followed by two iterative consensus rounds. The Delphi method was selected for its recognized capacity to structure expert judgment in areas characterized by conceptual complexity or limited empirical stabilization [26-29]. It has been widely used in healthcare research to develop and validate quality indicators [4,5,30].

The design, conduct, and reporting followed the CREDES (Conducting and Reporting Delphi Studies) recommendations [31]. Data were collected anonymously through online questionnaires between April and August 2025. The objective was to establish consensus on indicators capable of discriminating advanced levels of clinical competence in musculoskeletal physiotherapy.

### Participants and recruitment

The panel was composed exclusively of physiotherapists. This choice was deliberate and aligned with the study objective: to discriminate levels of clinical competence within the profession. In Delphi studies, alignment between panel composition and the field of practice under investigation is essential to ensure content validity, particularly when indicators concern situated clinical reasoning processes.

Recruitment combined convenience sampling, snowballing, and broad dissemination via physiotherapy schools, professional associations, continuing education networks, and social media.

An expertise assessment grid was developed to ensure a highly qualified and heterogeneous panel. The grid, reviewed and adjusted by three senior experts, provided a maximum score of 55 points across five domains: Academic training (10 points); Postgraduate professional training (20 points); Clinical experience within the previous ten years (5 points); Scholarly contributions (10 points); Professional recognition and leadership (10 points) (see Supplementary Material).

Each candidate was independently evaluated by two assessors. Discrepancies exceeding 15% within a domain were resolved through structured discussion to harmonize scoring. A score strictly above 30 points was required for inclusion, in order to ensure a sufficiently high level of expertise while maintaining adequate diversity within the panel.

The target panel size (30 participants) was consistent with methodological recommendations for exploratory Delphi studies in healthcare [27,28,30]. Final selection balanced high overall expertise with representation across expertise domains to maximize diversity while maintaining scientific rigour.

### Exploratory pre-round

A preliminary exploratory phase was conducted prior to the consensus rounds. The inclusion of a pre-round was justified by the absence of stabilized conceptual frameworks or empirically validated indicators specifically targeting discrimination of advanced levels of clinical competence in musculoskeletal physiotherapy [29].

The pre-round was administered via LimeSurvey® between 14 April and 15 May 2025. Participants received a theoretical framework clarifying the concepts of “indicator” and “advanced levels of clinical competence” as well as a methodological overview of the Delphi process (see Supplementary Material).

The questionnaire included: Six open-ended questions inviting experts to propose indicators and thirty-nine closed-ended items derived from the systematic review by Braybrooke et al. [21], rated on a six-point Likert scale.

The integration of literature-derived indicators served a dual purpose: to examine their perceived discriminative relevance and to support deductive structuring during thematic analysis.

Qualitative responses were analyzed inductively. Literature-based items informed deductive coding [4,7]. Two researchers independently conducted the analysis, and disagreements were resolved through discussion. The resulting synthesis generated a consolidated list of candidate indicators submitted to Round 1.

### Consensus rounds

Two Delphi rounds were conducted between June and August 2025. Each round remained open for four weeks and was separated by a two-week analysis period. Experts rated each indicator across three dimensions:

1. Relevance for discriminating advanced clinical competence
2. Clarity of formulation
3. Adequacy of proposed measurement tools

Ratings were provided using six-point Likert scales (1 = not appropriate at all; 6 = entirely appropriate).

Between rounds, quantitative analysis identified items reaching consensus. Qualitative comments were used to reformulate, merge, or generate additional indicators. Only items not reaching consensus were resubmitted in Round 2.

### Definition of consensus

In the present study, inclusion consensus was defined a priori as ≥75% of ratings in the upper categories (5 or 6). Disagreement was defined as ≥75% of ratings in the lower categories (1 or 2), resulting in exclusion.

The 75% threshold was selected according to the literature to ensure a focused set of indicators demonstrating strong discriminative potential, while preserving exploratory sensitivity in a conceptually complex domain literature [27,28]. Items not reaching either agreement or disagreement thresholds were classified as uncertain and resubmitted in the subsequent round.

### Data management and analysis

Qualitative analyses were conducted using QCAmap software. Quantitative analyses were performed using Microsoft Excel® (Microsoft 365, PC version). All data were stored locally on an encrypted external hard drive (AES-256 encryption), password-protected, and accessible only to authorized members of the research team.

### Ethical considerations

The study was conducted as a non-interventional investigation of professional practices and did not fall within the scope of research involving human subjects under French Public Health regulations (Article L1121-1).

Participation was voluntary. Informed consent was obtained prior to inclusion. Data were pseudonymized at the analysis stage and processed in compliance with the General Data Protection Regulation (GDPR). Analyses were conducted on aggregated data.

## RESULTS

### Participants and panel stability

Seventy-seven individuals were initially recruited (35 through convenience sampling, 9 via snowballing, and 33 through broad dissemination). Thirty-one were excluded (30 due to an expertise score ≤30 and one non-physiotherapist), leaving 46 eligible participants. Of these, 36 provided informed consent. Thirty-two completed the pre-round and were included in the study; 29 participated in Round 1 and 27 in Round 2, corresponding to a retention rate of 84.4% between the pre-round and the final round. Attrition was primarily due to time constraints within the allocated response periods.(Figure 1).

**Figure 1.**
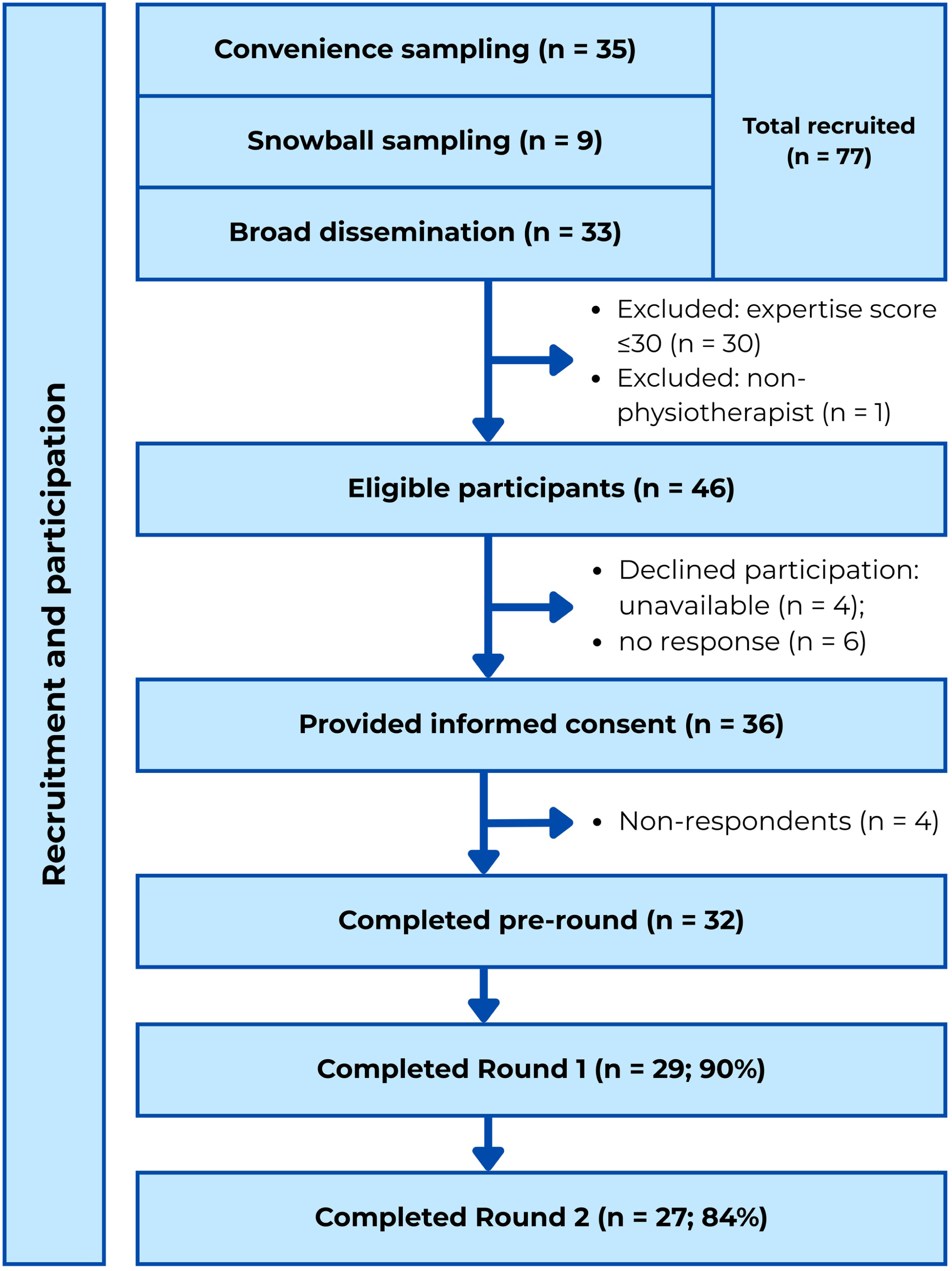
Flowchart of participant recruitment and participation throughout the study.

Participants demonstrated high levels of expertise. Half held a PhD; 12 had completed advanced postgraduate musculoskeletal training; 16 reported at least 10 years of clinical practice within the previous decade; 7 had authored more than ten international publications; and 3 held institutional or international leadership positions. The overall expertise score remained stable across rounds (40.6 ± 6.7 to 41.4 ± 6.8), indicating panel consistency (Table 1).

**Table 1.** Characteristics of the Delphi panel across study rounds.

|  | **Pre-round** | **Round 1** | **Round 2** |
| --- | --- | --- | --- |
| **Participants, n (%)** | 32 (100%) | 29 (90%) | 27 (84%) |
| **Countries of practice (n)** | United Kingdom (1)<br>Belgium (8)<br>France (18)<br>Quebec (2)<br>Switzerland (2)<br>Australia (1) | United Kingdom (0)<br>Belgium (8)<br>France (16)<br>Quebec (2)<br>Switzerland (2)<br>Australia (1) | United Kingdom (0)<br>Belgium (7)<br>France (15)<br>Quebec (2)<br>Switzerland (2)<br>Australia (1) |
| **Expertise score, mean ± SD** |  |  |  |
| **Total (/55)** | **40.6 ± 6.7** | **40.9 ± 6.8** | **41.4 ± 6.8** |
| Academic level (/10) | 7.9 ± 2.5 | 7.9 ± 2.5 | 8.3 ± 2.2 |
| Postgraduate MSK training (/20) | 17.2 ± 2.7 | 17.3 ± 2.7 | 17.3 ± 2.8 |
| Clinical experience (/5) | 4.3 ± 0.9 | 4.3 ± 0.9 | 4.3 ± 1 |
| Publications (/10) | 5.9 ± 3.5 | 6 ± 3.5 | 6.2 ± 3.6 |
| Leadership (/10) | 5.2 ± 2.4 | 5.4 ± 2.4 | 5.3 ± 2.5 |

*SD: Standard Deviation*

### Exploratory pre-round

Thirty-two experts completed the pre-round questionnaire. The combined inductive and deductive analysis generated 49 candidate indicators, categorized into five domains: outcomes (n = 8), process (n = 28), access and care pathways (n = 8), professional development (n = 2), and safety and regulatory framework (n = 3). These indicators were submitted to Round 1 (Figure 2).

**Figure 2.**
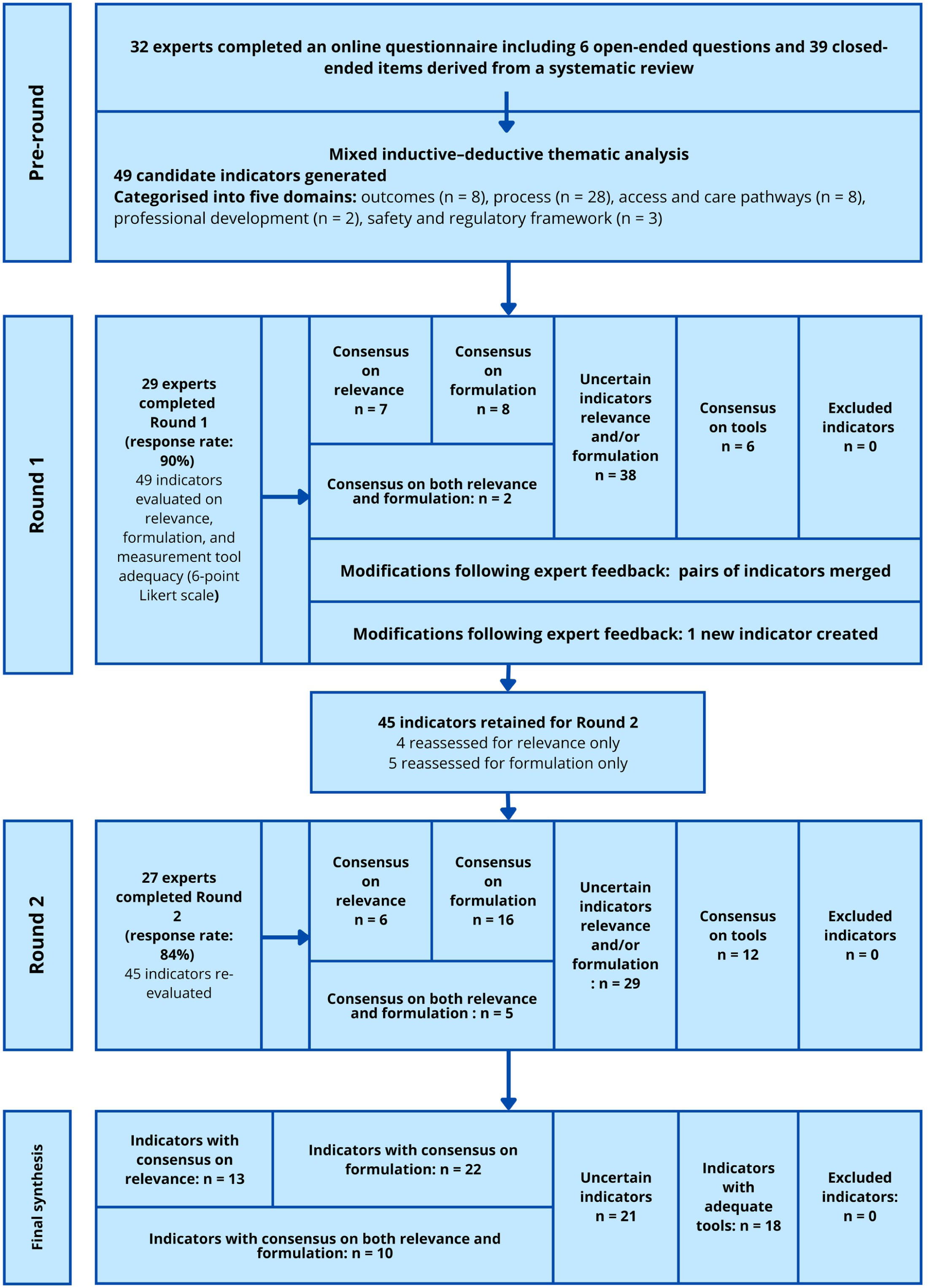
Delphi process and selection of clinical competence indicators across study phases.

### Round 1

Twenty-nine experts (90.6% of pre-round participants) completed Round 1.

Among the 49 indicators, 7 reached consensus on relevance and 8 on formulation, with 2 achieving simultaneous consensus on both dimensions. Six indicators reached consensus regarding the adequacy of proposed measurement tools. No indicator met criteria for exclusion through negative consensus. The majority of indicators (n = 38) were classified as uncertain on at least one evaluated dimension.

Following qualitative analysis of expert comments, two indicators that had achieved simultaneous consensus were removed from further evaluation. Three pairs of indicators were merged, and one new indicator was generated, resulting in 45 indicators being carried forward to Round 2.

### Round 2

Twenty-seven experts (84.4%) participated in Round 2.

Of the 45 indicators reassessed, 6 reached consensus on relevance and 16 on formulation, including 5 that achieved simultaneous consensus on both dimensions. Twelve indicators reached consensus regarding adequacy of measurement tools. No indicator reached negative consensus for exclusion.

### Final classification of indicators

After two rounds, 10 indicators achieved simultaneous consensus on relevance and formulation, 3 on relevance only, and 12 on formulation only. Twenty-one indicators remained uncertain.

The 13 indicators achieving consensus on relevance primarily reflected dimensions related to advanced clinical reasoning and management of complexity. They concerned dynamic adaptation of management plans during follow-up, prioritization of competing hypotheses, anticipation of clinical trajectories, decision-making under uncertainty, prognostic reasoning integrating biopsychosocial factors, appropriate referral of complex cases, and the management and integration of adverse events.

Notably, only one outcome-oriented indicator—clinical performance weighted by case complexity—reached strong consensus. In contrast, global outcome measures such as pain reduction, functional improvement, patient satisfaction, treatment adherence, and recurrence rates did not achieve consensus as discriminative markers of advanced clinical competence.

A detailed presentation of indicator scores and associated measurement tools is provided in Table 2.

**Table 2.**
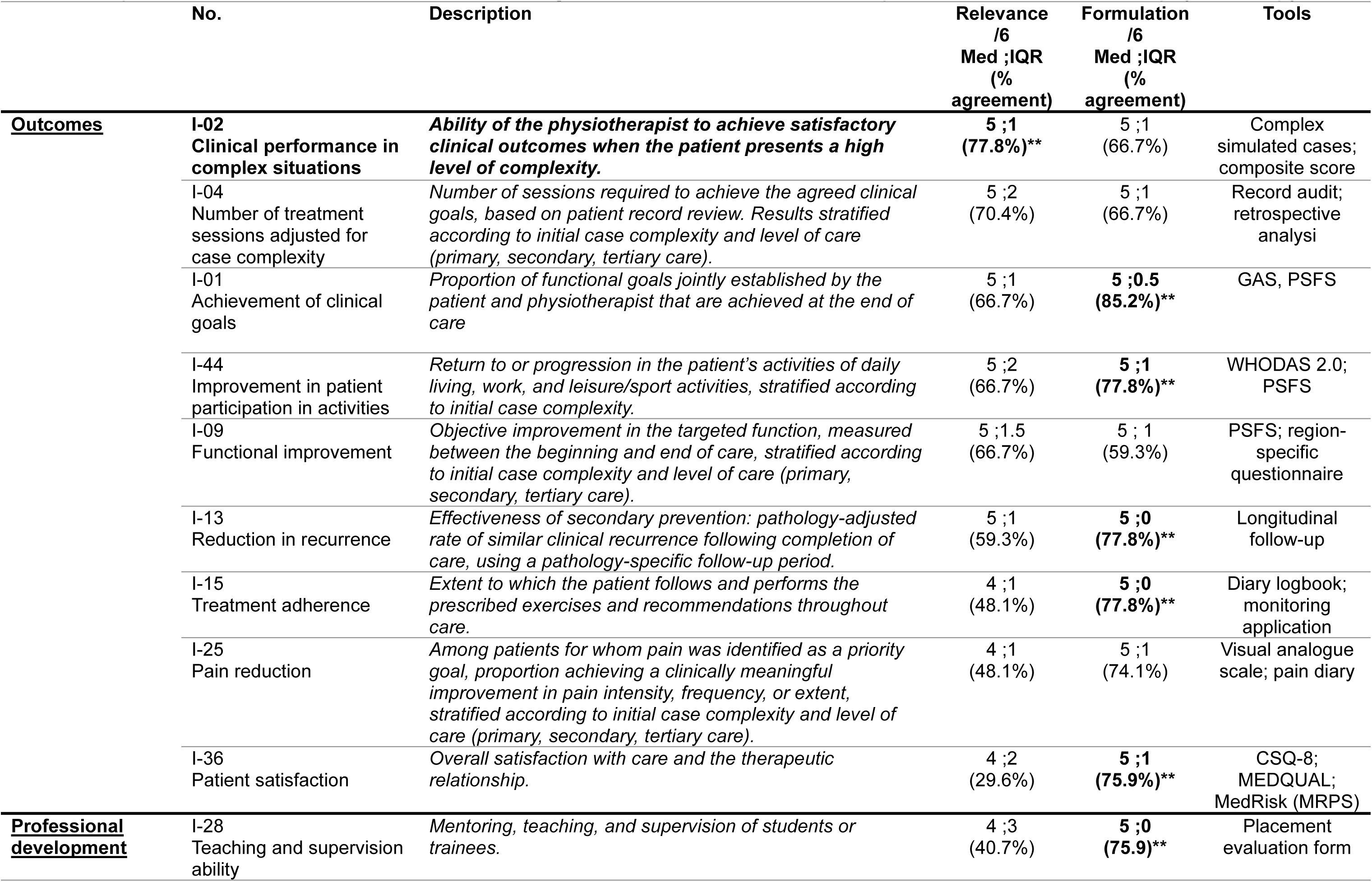
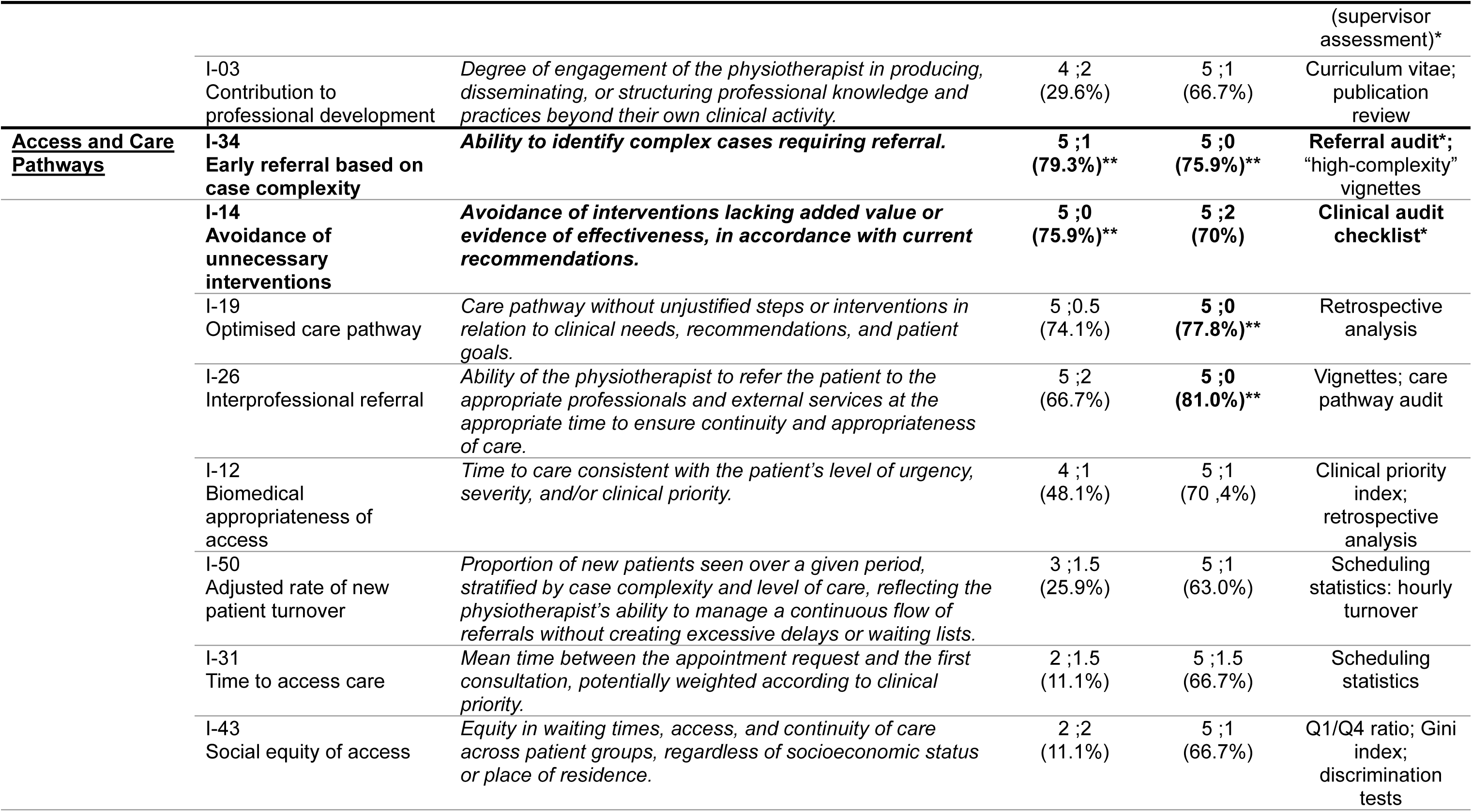
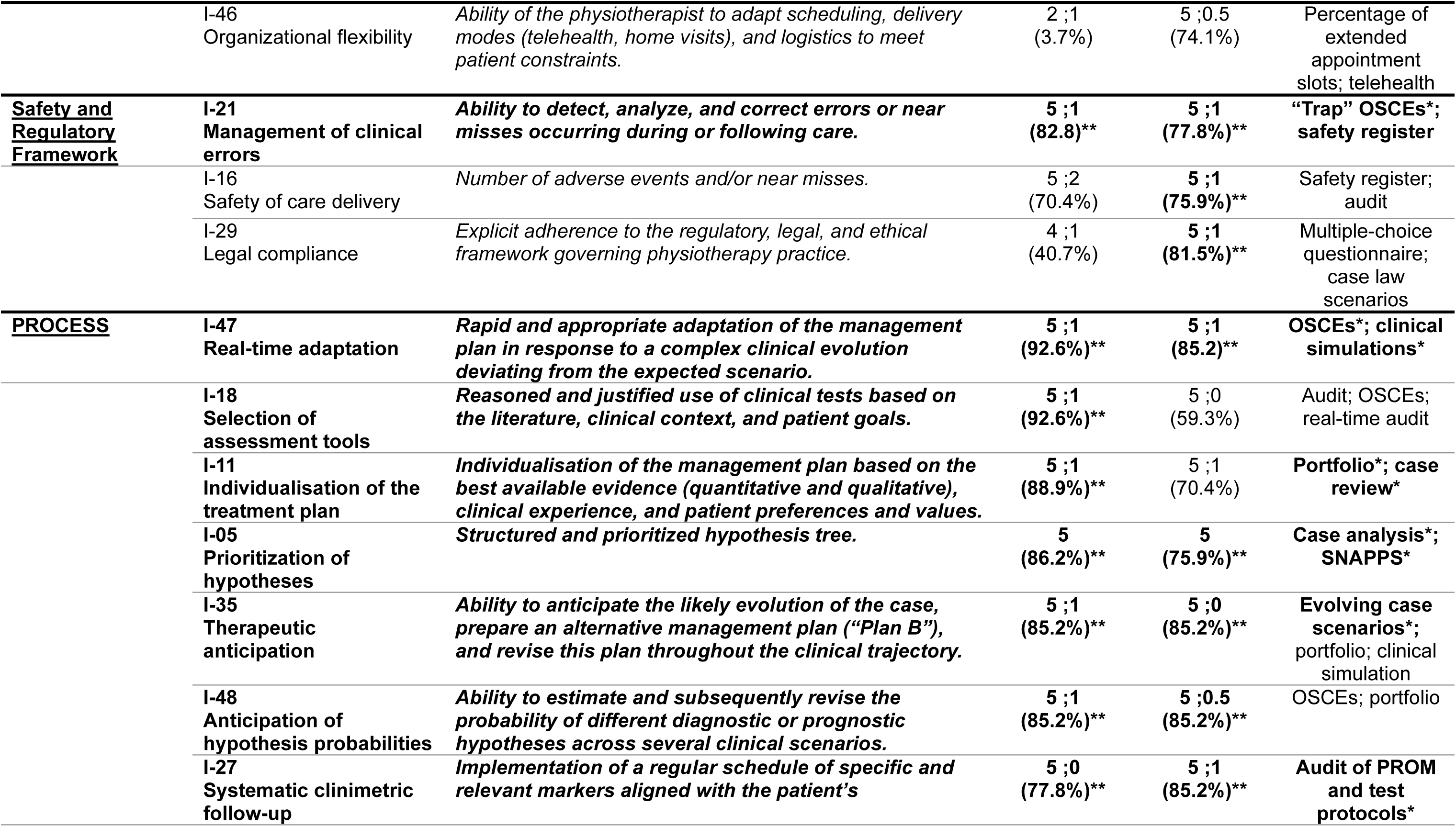
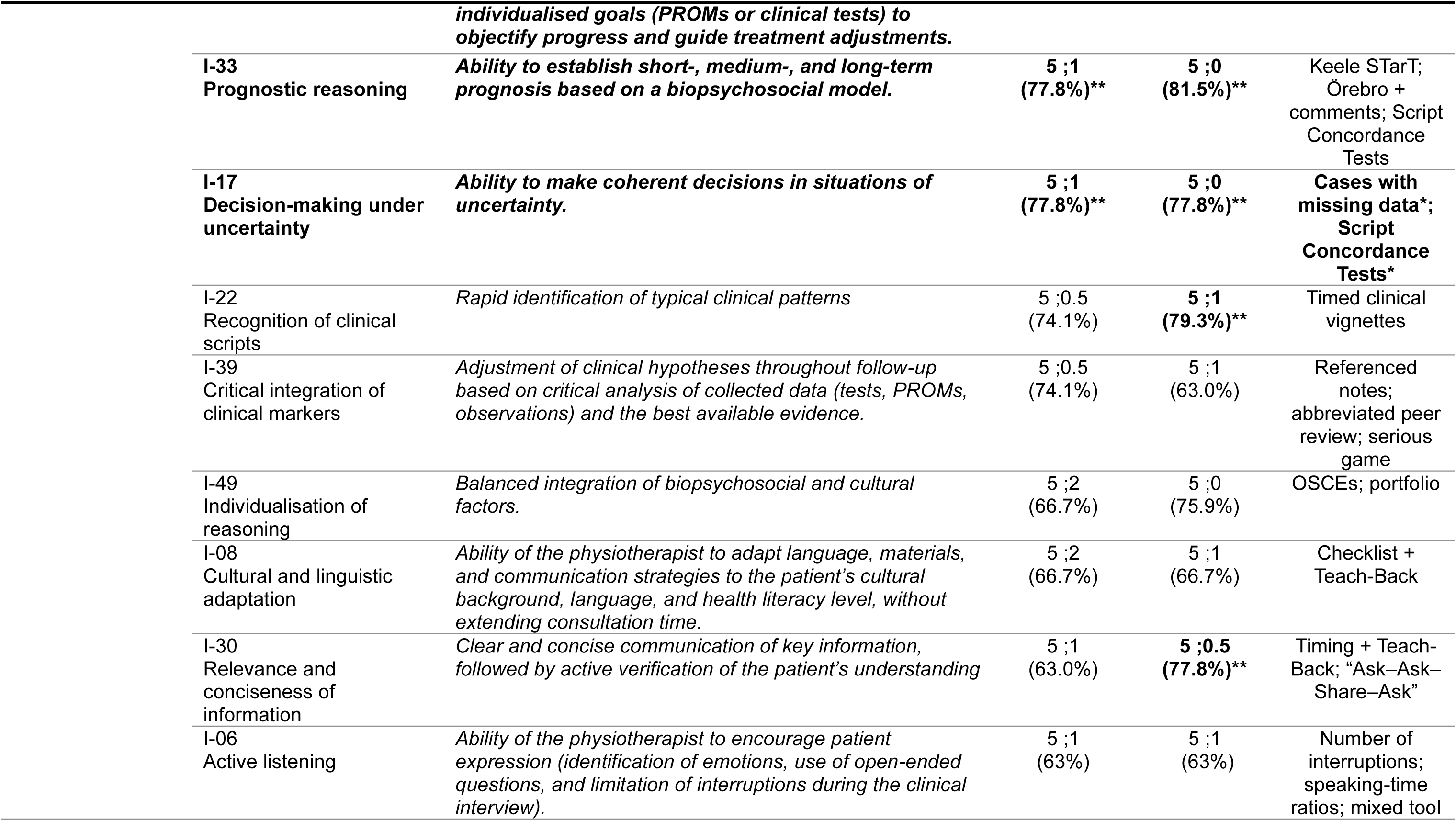
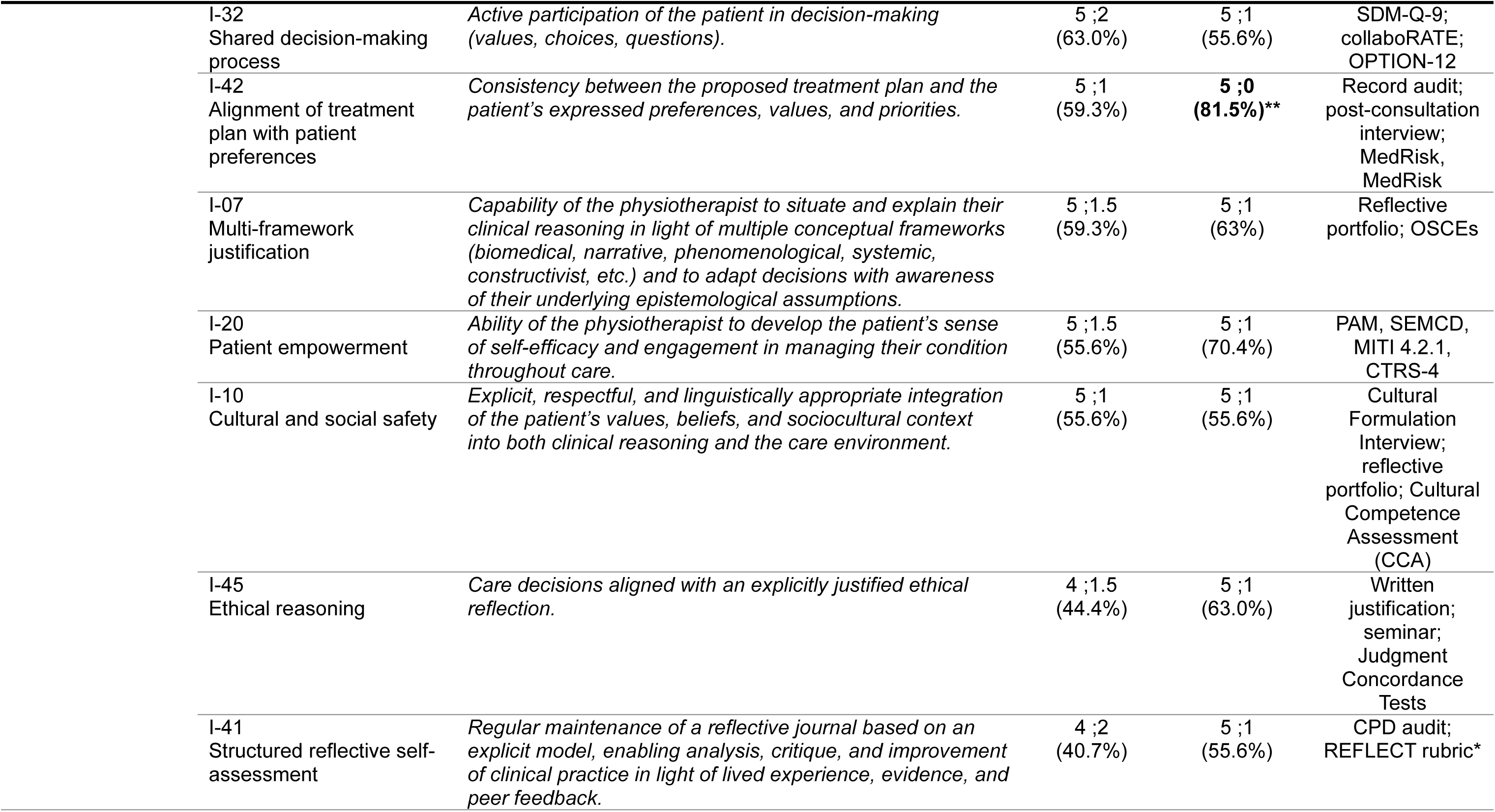
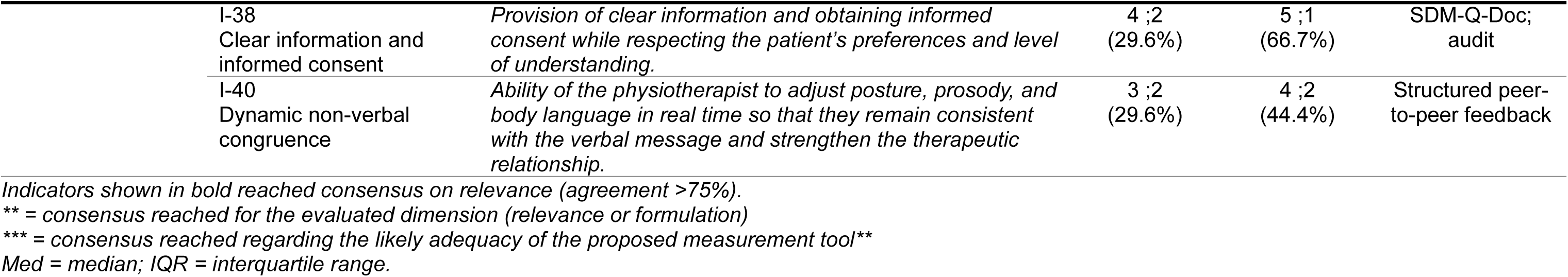
Proposed indicators evaluated for discriminating advanced levels of clinical competence in musculoskeletal physiotherapy.

## DISCUSSION

This study identified 13 indicators reaching consensus on relevance for discriminating advanced levels of clinical competence in musculoskeletal physiotherapy. These indicators predominantly reflect reasoning processes and complexity management rather than standardized outcomes.

The present study aimed to discriminate advanced levels of individual clinical competence, rather than to measure performance or to evaluate overall quality of care. Although Donabedian’s structure–process–outcome model provides a useful framework for organizing indicators, it does not in itself establish their ability to discriminate levels of expertise. The present analysis does not conflate clinical competence with quality of care; it identifies, within this framework, those indicators that more directly reflect processes characteristic of individual expertise.

### Structure and robustness of the indicator corpus

This study relied on a stable and highly engaged Delphi panel across all phases. The consistency of mean expertise scores between rounds, combined with high retention despite the protocol’s duration and demands, supports the robustness of the consensus and the relevance of the research question. Geographical diversity and complementary clinical, academic, and scientific profiles further strengthen internal validity, consistent with methodological recommendations for healthcare Delphi studies [27,30].

The exploratory pre-round generated a deliberately broad corpus of 49 indicators, avoiding premature restriction of the field. The subsequent two-round process followed an iterative evaluation consistent with established standards for clinical indicator development [5,6].

The predefined disagreement threshold (≥75% of ratings 1 or 2 on a six-point scale) was intentionally stringent and resulted in no formal exclusions and a final corpus containing a substantial proportion of uncertain items. Alternative strategies—such as lower thresholds, exclusion based on median values, or combined dispersion and central tendency criteria— were considered but not retained in order to preserve exploratory sensitivity in a domain where discriminative indicators remain insufficiently stabilized. The findings should therefore be interpreted as a structured clarification rather than a definitive indicator set.

Although the retained indicators span all dimensions of Donabedian’s model, this coverage does not guarantee discriminative value for advanced individual competence. This limitation is particularly salient for outcome indicators. Measures such as pain reduction, patient satisfaction, treatment adherence, or number of sessions are strongly influenced by case complexity, biopsychosocial determinants, and organizational or socioeconomic constraints largely independent of practitioner action. Without explicit stratification, attributing these outcomes solely to individual competence remains methodologically fragile.

By contrast, indicators centered on reasoning processes, complexity management, and decision-making under uncertainty are conceptually closer to markers of individual expertise, as they more directly reflect practitioner-level cognitive and adaptive processes and are comparatively less sensitive to contextual confounding.

### Defining the scope of clinical expertise

Although the exploratory pre-round generated indicators related to continuing professional development, research engagement, or structured reflective practice (e.g., I-28, I-03, I-41), these were not retained as discriminative of advanced clinical competence. Their exclusion does not diminish their professional value; rather, it reflects a deliberate refocusing on clinical expertise as expressed in patient care situations.

This result is particularly informative given the heterogeneous composition of the panel, which included academic and institutional experts. The convergence toward a circumscribed definition of clinical competence—distinct from broader roles such as teaching, research, or leadership—suggests a clear conceptual boundary between core clinical expertise and peripheral professional contributions, thereby strengthening the construct validity of the study.

### Non-discriminative indicators: distinguishing expected from advanced competence

A central finding of this study is the implicit hierarchy drawn by the panel between indicators documenting quality of care and those genuinely discriminative of advanced clinical competence. Indicators traditionally considered core to clinical practice—such as pain reduction (I-25), functional improvement (I-09), patient satisfaction (I-36), treatment adherence (I-15), shared decision-making (I-32), safety of care delivery (I-16), and recurrence reduction (I-13)—did not reach the consensus threshold required to characterize advanced competence.

While these outcomes remain essential for guiding management and demonstrating overall effectiveness, the panel considered them insufficient, in isolation, to differentiate a highly competent practitioner from one meeting expected professional standards. Their non-selection therefore does not diminish their clinical importance but suggests that they reflect a foundational level of competence rather than markers of advanced levels of clinical competence. This distinction clarifies the transition, from the experts’ perspective, between normative competence and genuinely discriminative advanced clinical performance.

### Contextual bias and validity limitations of outcome indicators

Beyond distinctions between competence levels, experts highlighted structural limitations affecting the validity of certain indicators, particularly outcome-based measures. Observed patient improvements were considered highly dependent on factors extrinsic to the intervention—such as individual characteristics, comorbidities, psychosocial determinants, and organizational constraints—many of which lie beyond the practitioner’s direct control. Recurrence rate illustrates this contextual bias. Several experts noted that the most complex cases, or those responding poorly to generalist management, are more frequently referred to highly experienced practitioners (see Supplementary Materials). In this context, a recurrence rate exceeding population averages may reflect case-mix concentration rather than deficient competence. Such observations underscore the methodological fragility of unadjusted outcome indicators for discriminating advanced clinical expertise. Comparable reservations were expressed regarding access-related indicators, including waiting times, organizational flexibility, and optimized care pathways. Although essential for evaluating system-level performance, these measures were regarded as strongly shaped by structural and territorial constraints and therefore insufficiently specific to individual clinical competence.

### Discriminative indicators and the specificity of advanced levels of clinical competence

Conversely, several indicators were judged highly discriminative of advanced levels of clinical competence. These share a common feature: they describe reasoning and adaptive processes in complex situations rather than standardized outcomes. The indicator “clinical performance in complex situations” (I-02) occupies a central position. It achieved strong consensus on relevance but not on formulation, indicating broad agreement on its conceptual importance alongside uncertainty regarding operationalization. Importantly, I-02 does not refer to attainment of a specific outcome, but to the ability to achieve meaningful results in contexts marked by uncertainty, multimorbidity, and non-linear trajectories—dimensions insufficiently formalized in conventional evaluation frameworks. The discrepancy likely reflects both the conceptual novelty of this indicator and heterogeneity within the panel regarding feasible methods of objectification.

Similarly, therapeutic anticipation (I-35), prioritization of hypotheses (I-05), and decision-making under uncertainty (I-17) characterize an advanced reasoning stance grounded in metacognitive regulation and dynamic adaptation. The inclusion of clinical error management (I-21) further reinforces this profile, valuing not error avoidance but the capacity to detect, analyze, and integrate errors as a learning mechanism, consistent with contemporary perspectives on patient safety and expertise development.

Notably, only one outcome-oriented indicator achieved strong consensus: clinical performance weighted by case complexity. Unlike traditional outcome measures, this indicator evaluates performance relative to contextual difficulty rather than in absolute terms. This approach aligns with recommendations advocating contextualized, change-sensitive, and clinically meaningful measurement beyond simple aggregation of PROMs and PREMs [32-34] and converges with recent work by Braybrooke et al, emphasizing the importance of accounting for complexity in musculoskeletal primary care.

### Positioning within the existing literature

The use of indicators carries a well-documented risk of gaming, whereby practices may shift toward optimization of measured variables once they become explicit targets, potentially at the expense of clinical goals [4,35,36]. This risk is particularly salient in normative or summative contexts. By contrast, audit and feedback interventions are more effective when embedded within contextualized, formative reflection rather than performance control [9,10]. The indicators proposed here should therefore be considered dynamic reference points integrated into reflective processes and periodically re-evaluated to preserve their discriminative value.

The present findings do not contradict the systematic review by Braybrooke et al, but reinterpret it through the lens of discriminating advanced competence. Although their work robustly identified quality indicators for musculoskeletal primary care, it did not explicitly differentiate competence levels, a limitation reflected in modest scores for discriminative capacity within Domain 4 of the AIRE instrument. By focusing directly on this dimension, the present study yields a more restricted yet more targeted set of indicators, potentially more operational for supporting development among already competent practitioners. Traditional quality indicators remain essential for ensuring minimum safety and performance standards; however, the indicators identified here highlight dimensions of advanced clinical expertise insufficiently operationalized in the literature.

These findings can also be situated within international frameworks describing levels of physiotherapy practice. Noblet, Newsham-West, Dean, and Rushton [37] emphasize clarifying distinctions between specialist, advanced, and expert roles to enhance transparency and patient safety. Contemporary multiprofessional frameworks conceptualize advanced practice as integrating multiple pillars—clinical expertise, leadership/management, education, and research [38-40]. A major contribution of the present study lies in specifying what discriminates high-level clinical competence independently of these additional pillars. The absence of consensus for indicators related to research, professional development, or structured reflection reflects a deliberate focus on the clinical pillar, consistent with Noblet et al.’s distinction between clinical specialization and broader advanced practice roles.

A similar interpretation applies to collaborative practice. Although interprofessional coordination is central to healthcare quality, it is often positioned within leadership and organizational domains extending beyond the therapist–patient dyad. The limited discriminative value attributed to access- and pathway-related indicators suggests that, while essential at system level, these dimensions do not constitute specific markers of individual clinical expertise.

Overall, the findings represent a targeted operationalization of advanced-level markers within the clinical pillar, defined by management of complexity, uncertainty, and dynamic adaptation in patient care.

### Synthesis of key findings

Consistent with Delphi methodology, the present findings reflect structured expert consensus on the relevance and formulation of candidate indicators rather than empirical validation of their discriminative power. These indicators should therefore be considered provisional and require further testing of reliability, feasibility, sensitivity to change, and discriminative capacity before operational use.

Overall, advanced clinical competence in musculoskeletal physiotherapy appears to be distinguished less by superior standardized outcomes than by the ability to manage complexity, uncertainty, and non-linear trajectories. Although several proposed indicators relate to outcomes (e.g., pain, satisfaction, number of sessions), their attribution to individual competence remains limited due to contextual, organizational, socioeconomic, and case-mix influences.

By contrast, the indicators achieving strongest consensus emphasize advanced reasoning processes, therapeutic anticipation, adaptive decision-making, integration of error as a learning mechanism, and performance contextualized by case complexity. By hierarchizing indicators according to discriminative value and prioritizing those more directly linked to clinical action, this study proposes a framework that may support more nuanced and development-oriented feedback systems.

### Limitations

Including multiple stakeholder groups or distinct cohorts would have broadened perspectives. However, in the absence of prior stabilized indicators, such an approach was not methodologically feasible at this stage. Broader stakeholder involvement should therefore be considered in subsequent phases, once the indicators have been further clarified.

The pre-round combined open-ended questions, a shared theoretical framework, and literature-derived items to reduce conceptual heterogeneity. Although consistent with the exploratory objective, this strategy may have introduced a framing effect, potentially orienting expert responses and limiting fully inductive exploration.

The number of indicators generated exceeded expectations and extended the duration of the process. While attrition remained moderate, the breadth of the corpus may have limited stabilization. A third round focused on items close to the consensus threshold might have facilitated further refinement. However, given the risk of additional attrition and the intention to preserve a distinction between robust consensus and fragile agreement, the decision to conclude after two rounds was deliberate. The substantial proportion of uncertain items likely reflects persistent heterogeneity of expert judgment in a conceptually complex and insufficiently stabilized domain rather than a methodological shortcoming.

The study did not aim to validate measurement instruments, and the panel was not constituted for that purpose. Proposed tools should therefore be interpreted as exploratory suggestions rather than implementation-ready recommendations. Similarly, feasibility was not a primary objective. Participants were encouraged to identify conceptually relevant indicators beyond current organizational constraints, yet several noted potential implementation challenges. This tension between conceptual relevance and practical feasibility remains to be addressed in future validation studies.

Finally, no established reference framework currently defines indicators characteristic of a “competent” baseline level of practice. Some disagreements reflected the perception that certain items represented expected entry-level competence rather than advanced levels of clinical competence. The absence of validated intermediate reference points limits hierarchical differentiation and underscores the need for future research dedicated to defining and validating indicators associated with a competent level of practice. Such work will inevitably remain context-dependent, as competence standards evolve across healthcare systems and professional environments.

### Future directions

Three main directions emerge.

First, developing a clinical complexity stratification tool appears essential to operationalize performance weighted by case complexity and ensure fair comparison across practitioners and contexts.

Second, the selected indicators require empirical validation, including assessment of feasibility within routine practice and evaluation of their reliability, validity, sensitivity to change, and true discriminative capacity.

Third, indicators related to other professional dimensions—such as leadership, collaboration, education, and research—should be explored through frameworks distinct from those mobilized here, as the present study deliberately focused on the clinical pillar. Finally, it should be acknowledged that indicators reflect underlying value judgments regarding what constitutes advanced competence. The emphasis placed on reasoning, uncertainty management, safety, and contextual adaptation corresponds to a situated conception of clinical expertise grounded in expert physiotherapists’ perspectives. Broader stakeholder involvement, including patients and other health professions, would allow expansion of the evaluative scope and examination of the ethical and organizational implications of such indicators.

## CONCLUSIONS

To our knowledge, this is the first study to identify indicators explicitly designed to discriminate advanced levels of clinical competence in musculoskeletal physiotherapy. The findings suggest that advanced levels of clinical competence is defined less by standardized outcomes than by metacognitive reasoning, adaptive decision-making, integration of error as a learning mechanism, and engagement with clinical uncertainty. These indicators offer potential leverage points for deliberate practice and competence-oriented feedback, such as within structured clinical supervision integrating case-complexity stratification. Further research should refine their operationalization across competence levels, evaluate their feasibility and instrumental validity in clinical settings, and incorporate broader stakeholder perspectives—particularly those of patients—to consolidate a more comprehensive framework for evaluating clinical expertise.

## DATA AVAILABILITY STATEMENT

The data that support the findings of this study are available from the corresponding author upon reasonable request.

## ACKNOWLEDGEMENTS

The authors sincerely thank all experts who participated in the Delphi process, whether across all phases or at specific stages. Their engagement substantially strengthened the quality and depth of the findings. The authors also acknowledge the professionals who supported the initiative and expressed interest in the project.

## USE OF GENERATIVE ARTIFICIAL INTELLIGENCE

During the preparation of this manuscript, the authors used generative artificial intelligence tools to assist with formatting and to suggest alternative wordings. No artificial intelligence tool was used to generate data, to conduct or interpret the analysis, or to produce the scientific content of this work. The authors reviewed and edited all suggested text, and take full responsibility for the content of the publication.

## CREDIT AUTHOR STATEMENT

A. Mounier-Poulat: Conceptualization, Methodology, Investigation, Data curation, Formal analysis, Project administration, Visualization, Writing – original draft, Writing – review & editing.

R. Artico: Conceptualization, Methodology, Validation, Formal analysis, Supervision, Writing – review & editing.

N. Pinsault: Conceptualization, Supervision, Validation, Writing – review & editing.

## SUPPLEMENTARY MATERIALS

*These supplementary materials were produced in French, the working language of the Delphi panel, and are therefore not deposited with this preprint, in accordance with medRxiv language policy. They are available from the corresponding author on request*.

Supplementary Material 1. Theoretical framework

Supplementary Material 2. Methodological procedure

Supplementary Material 3. Expertise assessment grid

Supplementary Material 4. Complete indicator set

Supplementary Material 5. Pre-round questionnaire

Supplementary Material 6. Recruitment materials

## REFERENCES

1. Gill TK, Mittinty MM, March LM, Steinmetz JD, Culbreth GT, Cross M, Kopec JA, Woolf AD, Haile LM, Hagins H, et al. Global, regional, and national burden of other musculoskeletal disorders, 1990–2020, and projections to 2050: A systematic analysis of the Global Burden of Disease Study 2021. The Lancet Rheumatology. 2023;5:e670–e682. doi:10.1016/S2665-9913(23)00232-1

2. Wollersheim H, Hermens R, Hulscher M, Braspenning J, Ouwens M, Schouten J, Grol R. Clinical indicators: Development and applications. Netherlands Journal of Medicine. 2007;65:15–22.

3. Donabedian A. The quality of care: how can it be assessed? Journal of the American Medical Association. 1988;260:1743–1748. doi:10.1001/jama.260.12.1743

4. Campbell SM, Braspenning J, Hutchinson A, Marshall MN. Research methods used in developing and applying quality indicators in primary care. British Medical Journal. 2003;326:816–819. doi:10.1136/bmj.326.7393.816

5. Kötter T, Blozik E, Scherer M. Methods for the guideline-based development of quality indicators—A systematic review. Implementation Science. 2012;7:21. doi:10.1186/1748-5908-7-21

6. Mainz J. Defining and classifying clinical indicators for quality improvement. International Journal for Quality in Health Care. 2003;15:523–530. doi:10.1093/intqhc/mzg081

7. Mainz J. Developing evidence-based clinical indicators: A state of the art methods primer. International Journal for Quality in Health Care. 2003;15:i5–i11. doi:10.1093/intqhc/mzg084

8. Baker R, Camosso-Stefinovic J, Gillies C, Shaw EJ, Cheater F, Flottorp S, Robertson N. Tailored interventions to overcome identified barriers to change: Effects on professional practice and health care outcomes. Cochrane Database of Systematic Reviews. 2010;3:CD005470. doi:10.1002/14651858.CD005470.pub2

9. Foy R, Skrypak M, Alderson S, Ivers NM, McInerney B, Stoddart J, Ingham J, Keenan D. Revitalising audit and feedback to improve patient care. British Medical Journal. 2020;368:m213. doi:10.1136/bmj.m213

10. Ivers N, Jamtvedt G, Flottorp S, Young JM, Odgaard-Jensen J, French SD, O’Brien MA, Johansen M, Grimshaw J, Oxman AD. Audit and feedback: Effects on professional practice and healthcare outcomes. Cochrane Database of Systematic Reviews. 2012;6:CD000259. doi:10.1002/14651858.CD000259.pub3

11. Ivers N, Yogasingam S, Lacroix M, Brown KA, Antony J, Soobiah C, Simeoni M, Willis TA, Crawshaw J, Antonopoulou V, et al. Audit and feedback: Effects on professional practice. Cochrane Database of Systematic Reviews. 2026;6:CD000259. doi:10.1002/14651858.CD000259.pub5

12. World Health Organization. Global report on quality of care. Geneva: World Health Organization; 2024. Available from: https://www.who.int/publications/i/item/9789240095458

13. Miller GE. The assessment of clinical skills/competence/performance. Academic Medicine. 1990;65:S63–S67. doi:10.1097/00001888-199009000-00045

14. Rethans JJ, Norcini JJ, Barón-Maldonado M, Blackmore D, Jolly BC, LaDuca T, Lew S, Page GG, Southgate LH. The relationship between competence and performance: Implications for assessing practice performance. Medical Education. 2002;36:901–909. doi:10.1046/j.1365-2923.2002.01316.x

15. Dreyfus HL, Dreyfus SE. Mind over machine: the power of human intuition and expertise in the era of the computer. New York: Free Press; 1986.

16. Epstein RM, Hundert EM. Defining and assessing professional competence. Journal of the American Medical Association. 2002;287:226–235. doi:10.1001/jama.287.2.226

17. Tardif J. L’évaluation des compétences: documenter le parcours de développement. Montréal: Chenelière Éducation; 2006.

18. Gruppen LD. Clinical reasoning and expertise. In: Norman GR, van der Vleuten CPM, Newble DI, editors. International handbook of research in medical education. Dordrecht: Springer; 2002. p. 295-314.

19. Ericsson KA, Krampe RT, Tesch-Römer C. The role of deliberate practice in the acquisition of expert performance. Psychological Review. 1993;100:363–406. doi:10.1037/0033-295X.100.3.363

20. Hattie J, Timperley H. The power of feedback. Review of Educational Research. 2007;77:81–112. doi:10.3102/003465430298487

21. Braybrooke A, Baraks K, Burgess R, Banerjee A, Hill JC. Quality indicators for the primary and community care of musculoskeletal conditions: A systematic review. Archives of Physical Medicine and Rehabilitation. 2025;106:459–472. doi:10.1016/j.apmr.2024.08.022

22. Ericsson KA, Pool R. Peak: secrets from the new science of expertise. Boston (MA): Houghton Mifflin Harcourt; 2016.

23. Jensen GM, Gwyer J, Shepard KF, Hack LM. Expert practice in physical therapy. Physical Therapy. 2000;80:28–43.

24. Mocke M, Unger M, Hanekom S. Validation of the World Health Organization Rehabilitation Competency Framework: An illustration using physiotherapy. Clinical Rehabilitation. 2025;39:88–98. doi:10.1177/02692155241300271

25. World Health Organization. Rehabilitation in health systems. Geneva: World Health Organization; 2020. Available from: https://iris.who.int/handle/10665/254506

26. Belton I, MacDonald A, Wright G, Hamlin I. Improving the practical application of the Delphi method in group-based judgment: A six-step prescription for a well-founded and defensible Delphi study. Technological Forecasting and Social Change. 2019;147:72–82. doi:10.1016/j.techfore.2019.07.002

27. Diamond IR, Grant RC, Feldman BM, Pencharz PB, Ling SC, Moore AM, Wales PW. Defining consensus: A systematic review recommends methodologic criteria for reporting of Delphi studies. Journal of Clinical Epidemiology. 2014;67:401–409. doi:10.1016/j.jclinepi.2013.12.002

28. Trevelyan EG, Robinson N. Delphi methodology in health research: how to do it? European Journal of Integrative Medicine. 2015;7:423–428. doi:10.1016/j.eujim.2015.07.002

29. Waggoner J, Carline JD, Durning SJ. Is there a consensus on consensus methodology? Academic Medicine. 2016;91:663–668. doi:10.1097/ACM.0000000000001092

30. Boulkedid R, Abdoul H, Loustau M, Sibony O, Alberti C. Using and reporting the Delphi method for selecting healthcare quality indicators: A systematic review. PLoS ONE. 2011;6:e20476. doi:10.1371/journal.pone.0020476

31. Jünger S, Payne SA, Brine J, Radbruch L, Brearley SG. Guidance on conducting and reporting Delphi studies (CREDES). Palliative Medicine. 2017;31:684–706. doi:10.1177/0269216317690685

32. Benson T. Why it is hard to use PROMs and PREMs in routine health and care. BMJ Open Quality. 2023;12:e002516. doi:10.1136/bmjoq-2023-002516

33. Cella D, Yount S, Rothrock N, Gershon R, Cook K, Reeve B, Ader D, Fries JF, Bruce B, Rose M. The Patient-Reported Outcomes Measurement Information System (PROMIS): Progress of an NIH roadmap cooperative group. Medical Care. 2007;45:S3–S11. doi:10.1097/01.mlr.0000258615.42478.55

34. NHS England. National PROMs consultation report. London: NHS England; 2017. Available from: https://www.england.nhs.uk/wp-content/uploads/2017/08/proms-consultation-report.pdf

35. Lilford RJ, Brown CA, Nicholl J. Use of process measures to monitor the quality of clinical practice. British Medical Journal. 2007;335:648–650. doi:10.1136/bmj.39317.641296.AD

36. Mannion R, Braithwaite J. Unintended consequences of performance measurement in healthcare: 20 salutary lessons from the English National Health Service. Internal Medicine Journal. 2012;42:569–574. doi:10.1111/j.1445-5994.2012.02766.x

37. Noblet T, Newsham-West R, Dean C, Rushton A. Defining our future: International use of contemporary levels of clinical practice will ensure the future of physiotherapy. Journal of Physiotherapy. 2025;71:219–224. doi:10.1016/j.jphys.2025.09.003

38. NHS England. The Centre for Advancing Practice: multi-professional framework for advanced practice in England (PRN01679). London: NHS England; 2025. Available from: https://advanced-practice.hee.nhs.uk/wp-content/uploads/sites/3/2025/01/Multi-professional-framework-for-advanced-practice-in-England.pdf

39. World Physiotherapy. Physiotherapist education framework. London: World Physiotherapy; 2021. Available from: https://world.physio/sites/default/files/2023-03/Physiotherapist-education-framework.pdf

40. World Physiotherapy. Policy statement: advanced physiotherapy practice. London: World Physiotherapy; 2023. Available from: https://world.physio/policy/ps-advanced-pt-practice

